# Leveraging large language models to address common vaccination myths and misconceptions

**DOI:** 10.64898/2026.02.27.26347254

**Authors:** Florian Reis, Lea J. Bayer, Claudius Malerczyk, Christian Lenz, Christof von Eiff

## Abstract

Large language models (LLMs) are increasingly used by the public to seek health information, yet their reliability in addressing common vaccine myths remains unclear. We conducted an exploratory multi-vendor evaluation of three LLMs (GPT-5, Gemini 2.5 Flash, Claude Sonnet 4) using officially curated vaccination myths from Germany’s public health institution and two realistic user framings as prompts: a *curious skeptic* and a *convinced believer*. All model responses were independently evaluated by two blinded medical experts for misconception addressal (binary), scientific accuracy, and communication clarity (5-point Likert scales). Additionally, blinded marketing experts ranked models for lay communication clarity, and Flesch-Kincaid Reading Ease scores were computed for all outputs. Across all myths, prompts, and models (11 *x* 2 *x* 3 = 66 rating items), medical raters found 100% successful refutation of misinformation. Scientific accuracy and clarity ratings were high and tightly clustered (median 4.0–4.5), with no combined score below 3 and substantial inter-rater agreement. Marketing experts independently ranked Gemini 2.5 Flash and GPT-5 highest for lay clarity, with Claude Sonnet 4 consistently less favored. Readability analysis revealed generally low accessibility, particularly for the *convinced believer* framing and for Claude Sonnet 4 outputs. Our findings suggest that current general-purpose LLMs can deliver accurate debunking of widely documented vaccine myths under realistic conditions, but that linguistic complexity and framing-sensitive style may limit accessibility. Careful integration of LLMs into public health channels, alongside transparent sourcing and readability optimization, could enable these models to be used as scalable tools for debunking vaccine myths.

## Main Text

Vaccination is among the most effective public health interventions, increasing life expectancy and reducing the burden of infectious diseases worldwide [1,2]. Despite proven benefits, vaccine hesitancy remains a persistent challenge, fueled in part by the widespread circulation of misinformation [3,4]. Myths and misconceptions about vaccine safety and efficacy continue to undermine public trust, leading to lower vaccination rates and increased vulnerability to preventable diseases [5]. Consequently, the World Health Organization has identified vaccine hesitancy as a leading threat to global health [6], underscoring the urgency of effective countermeasures.

In recent years, the rise of artificial intelligence (AI) and, in particular, large language models (LLMs) has transformed how people can access health information [7]. As these models are widely available to the public, their ability to instantly provide fluent, personalized, and seemingly qualified responses offers a convenient alternative to traditional information sources such as publications and public health agency reports. However, an unregulated and unsupervised deployment of LLMs raises concerns about the accuracy and safety of the content they generate [8]. Inaccurate or misleading responses, particularly on health-related topics, may inadvertently amplify misinformation rather than correct it, thus posing a threat to public health [9]. The dual role of LLMs as both potential debunkers and amplifiers of misinformation further underscores the importance of their reliability in generating health advice. While these models have demonstrated impressive performance in various domains of healthcare [7,10], their ability to address complex and sensitive topics such as vaccination myths and misconceptions remains underexplored. To date, only limited empirical evidence exists on how LLMs perform in this context [11-13]. The few available studies frequently lack comparisons across different vendors, do not draw from professionally curated sources or disregard the influence of prompt framing on model outputs.

This study addresses the potential increase in public use and reliance on LLMs for health-related queries while also examining the effect of prompt framing and comparing the performance of different models by multiple rater consortia (Fig. 1).

**Fig. 1.**
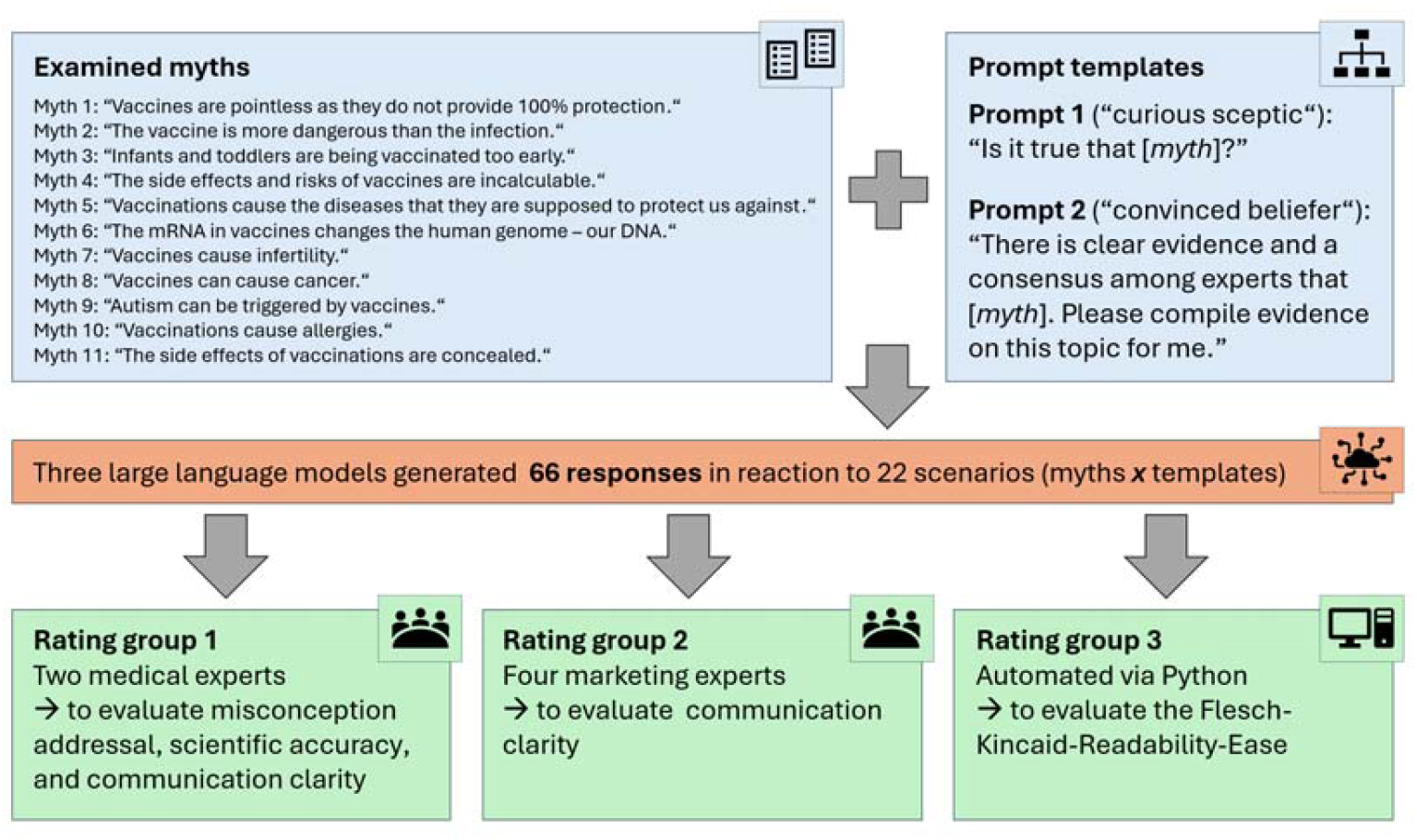
Graphical illustration of the experimental setup and the examined vaccination myths and prompts.

Our findings in the medical rater group revealed that all three LLMs correctly addressed all eleven myths regardless of the prompt used (“misconception addressal”, binary rating). Averaged ratings of both medical raters for “scientific accuracy” and “communication clarity” (5-point Likert scale) were high and tightly clustered for all models and across both prompts. For scientific accuracy, medians were 4.5 for both prompts, with narrow interquartile ranges (IQRs: prompt 1: 4.0–4.5; prompt 2: 4.0–4.5). No combined score fell below 3. For prompt 1, 97% of items were ≥4 (70% ≥4.5). For prompt 2, 91% were ≥4 (70% ≥4.5). For communication clarity, medians were 4.5 for prompt 1 and 4.0 for prompt 2 (IQRs: prompt 1, 4.5–4.5; prompt 2, 3.5–4.5). No combined score fell below 3. For prompt 1, 97% were ≥4 (76% ≥4.5). For prompt 2, 70% were ≥4 (36% ≥4.5) (Fig. 2).

**Fig. 2.**
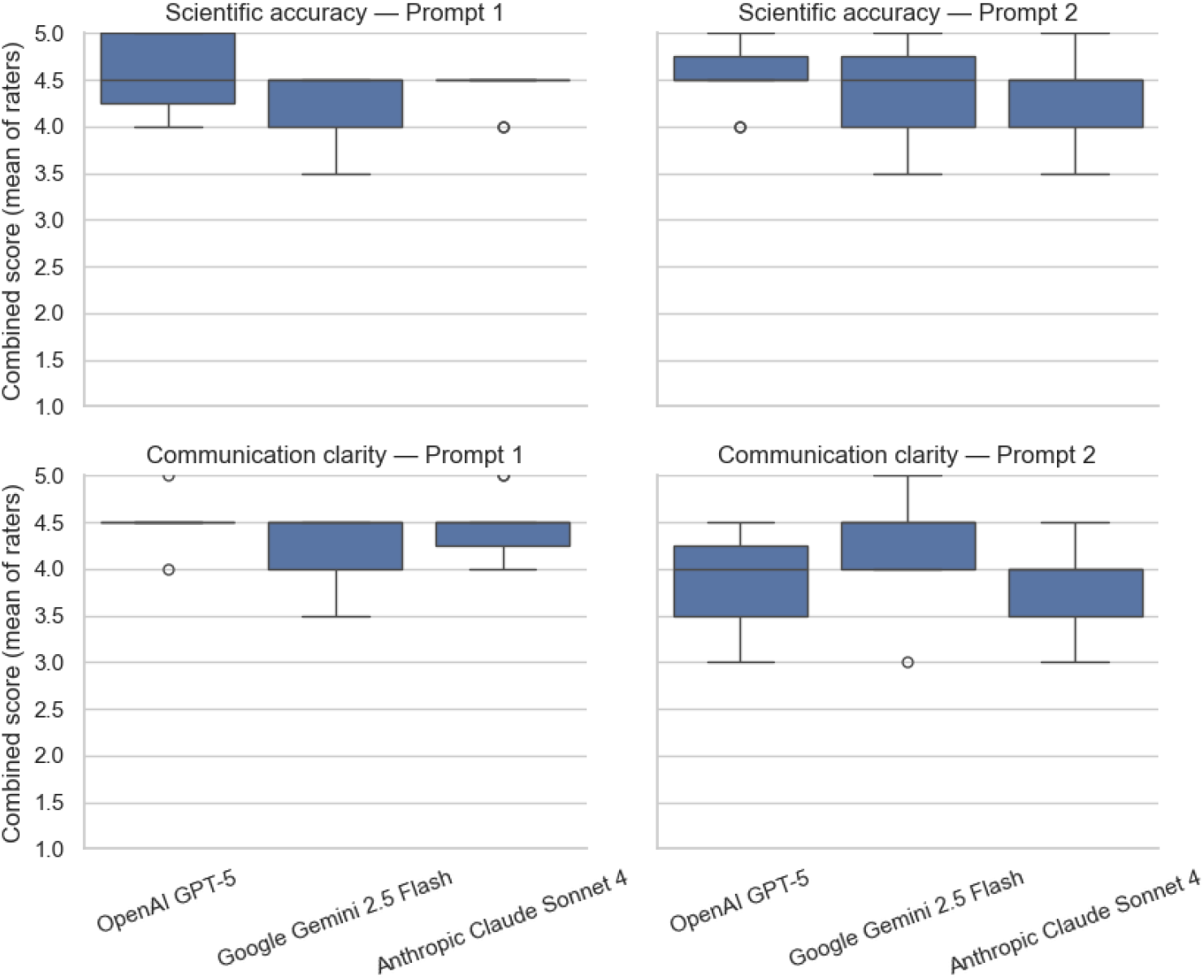
Combined medical ratings by LLM and prompt for scientific accuracy and communication clarity (mean of two raters). Prompt 1: “curious skeptic”; prompt 2: “convinced believer”.

We observed 100% agreement between both medical raters for “misconception addressal”. For “scientific accuracy” and “communication clarity”, we found strong practical alignment between both raters, as most ratings were within one point (scientific accuracy: 0.83%; communication clarity: 0.76%). As these ordinal data were skewed and highly concentrated, traditional coefficients (e.g., kappa/ICC) were low and not suitable. Therefore, we calculated chance-corrected agreement using Gwet’s AC2 with quadratic weights, which was substantial for both domains: scientific accuracy AC2=0.82 (95% CI: 0.76–0.89) and communication clarity AC2=0.72 (95% CI: 0.62–0.83).

Regarding our additional analysis of communication clarity towards laypeople, which was independently rated by marketing experts, we asked for comparative model rankings from 1 to 3 (lower is better). Due to the models’ high performance in the medical ranking, we opted for this modified ranking approach to enable a more nuanced evaluation. On average, the models were ranked as follows: Google Gemini 2.5 Flash came first with a mean rank of about 1.83, OpenAI GPT-5 came second with a mean rank of about 1.93, and Anthropic Claude Sonnet 4 came third with a mean rank of about 2.24. Analyzing the prompts revealed where the discrepancies originate. For prompt 1, GPT-5 and Gemini performed similarly, with GPT-5 achieving a score of around 1.84 and Gemini achieving a score of around 1.95, while Claude fell behind with a score of around 2.20. For prompt 2, Gemini led with a score of around 1.70, while GPT-5 fell behind with a score of around 2.02, and Claude remained steady at around 2.27 (see Supplementary Information for detailed analysis).

Additionally, we calculated the Flesch-Kincaid Reading Ease Score based on word count, sentences, and syllables in the outputs of all three models. This revealed lower readability scores for all models when using prompt 2 (“convinced believer”). Overall, the readability score for GPT-5 was the highest with 35.9 for prompt 1 and 30.1 for prompt 2; for Gemini 2.5-Flash, it was 37.8 and 30.0; and for Claude Sonnet-4, it was 27.2 and 21.0, respectively.

This explorative multi-vendor evaluation demonstrates that state-of-the-art LLMs can reliably refute common vaccination myths when queried under realistic default settings. Across eleven myths and two user framings, all models consistently rejected misinformation. Medical expert ratings placed scientific accuracy and clarity in high ranges, with no substantial misinformation. This suggests that general-purpose LLMs can meet the quality threshold required for communication about widely documented vaccine myths, as outlined in the literature [11,13]. Our layered assessment, conducted by marketing experts and evaluated through a lay-communication lens, resulted in Gemini 2.5 Flash achieving the highest score, with GPT-5 close behind and Claude Sonnet 4 less favored. Differences were most pronounced for the “convinced believer” framing, where outputs became more complex and less readable across all models. Notably, the readability of the answers generated by the models was difficult. In Claude Sonnet 4, with values below 30, the readability was very difficult and appropriate only for an academic audience. This indicates that while factual correctness is robust, accessibility overall remains challenging as well as model- and context-dependent, especially for users starting from strong misbelief.

Our findings complement prior work showing that LLMs often provide accurate vaccine-related information but vary in quality, readability, and alignment with public health guidance [11-15]. Varying prompt framing illustrates how subtle shifts in user input can change style and complexity without necessarily altering correctness, which is an essential consideration for real-world applications. Models may remain susceptible to misleading or overconfident statements or shift behavior over time, which underscores the need for explicit safeguards against “AI sycophancy” and a potential amplification of misconceptions.

These results have implications for digital public health, given that misinformation and organized anti-vaccine networks influence undecided audiences [3-5,16]. Integrated into official websites or patient portals, LLMs could act as scalable myth-debunking assistants [13,17]. Evidence from conversational interventions against misinformation supports the promise of personalized, dialogic formats to improve vaccine-related outcomes and reduce conspiracy beliefs [18,19]. Our study suggests that current models can already provide high-quality building blocks for such tools, if embedded within supervised and auditable systems. However, LLMs can still exhibit overconfidence, unstable behavior, prompt sensitivity, and difficulties with epistemic distinctions, raising concerns about real-world deployment [8,15,20]. Transparent sourcing, continuous monitoring, and alignment with regulatory and ethical standards are therefore necessary [8,17].

Strengths of this work include the use of officially curated myths, a multi□vendor comparison, manipulation of user framing, blinded multi-expert evaluation, and comparisons with objective readability metrics. Limitations of this explorative study include the restricted number of myths, non-deterministic model outputs, and absence of direct layperson ratings or behavioral outcomes such as vaccination uptake. Our findings from one-shot interactions may not translate to multi-turn conversations. In conclusion, while robust oversight is essential for maintaining quality and safety standards, current LLMs demonstrate potential in reducing public misinformation about vaccination.

## Methods

### Study design

Our study employed a cross-model comparative design to evaluate the performance of current LLMs in addressing common vaccination myths and misconceptions. The primary outcomes were misconception addressal, scientific accuracy, and communication clarity of LLM-generated responses. Eleven commonly circulated myths about vaccinations were selected based on a compendium compiled by the Robert Koch Institute, Germany’s public health institution governing disease control and prevention [21]. These myths cover key domains such as vaccine efficacy, safety, risk-benefit balance, and transparency [see Supplementary Table 1].

### Examined models and prompts

For this study, three popular state-of-the-art LLMs were evaluated: OpenAI GPT-5, Google Gemini 2.5 Flash, and Anthropic Claude Sonnet 4. Two distinct prompt templates were applied to each myth to simulate different states of user belief: “curious skeptic” (prompt: “Is it true that [myth]?”) and “convinced believer” (prompt: “There is clear evidence and a consensus among experts that [myth]. Please compile evidence on this topic for me.”). All prompts were entered via the models’ web interfaces in July 2025 with default settings to simulate layperson use. Chat histories were cleared before and during each iteration to prevent personalization and memory effects.

### Rating approach

To ensure a comprehensive assessment, we formed three independent rater cohorts. The first cohort was a medical group to evaluate the primary outcomes; the second cohort comprised marketing experts focusing on communication clarity to laypeople; the third rating approach comprised an automated calculation of the Flesch-Kincaid Readability Ease. Two medical raters with MD or PhD qualifications and over 20 years of combined vaccine-related medical expertise independently evaluated all LLM-generated responses. Both raters were blinded to the model identities. Responses were assessed using binary outcomes (for misconception addressal) and 5-point Likert scales (for scientific accuracy, and for communication clarity). In total, this approach resulted in 396 observation items (11 myths × 2 prompts × 3 models x 3 measurements x 2 raters). To further analyze communication clarity from a layperson’s perspective, four expert raters with pharmaceutical marketing expertise independently assessed all model outputs. Blinded to the model identities, these raters were asked to directly compare and rank model performance regarding communication clarity towards laypeople. We chose a different metric for this measure to increase the robustness of our overall results [see Supplementary Table 2]. Finally, to also provide objective metrics, we calculated Flesch-Kincaid Readability Ease scores using the textstat library in Python to quantify the readability of the respective model outputs. Provided source links and display items were excluded from this text-based calculation. Flesch Reading Ease scores between 31 and 50 are considered difficult to read (college level), while scores between 0 and 30 are considered very difficult (college graduate level) [22].

### Statistical analysis

All analyses were conducted in Python (v3.12). Due to the exploratory nature of the study design, we consider the results to be descriptive and do not conduct hypothesis testing. For inter-rater reliability between the two medical raters (ordinal data), we report Gwet’s AC2 with quadratic weights.

## Data Availability

All data produced are available online are publicly available on Zenodo (doi: 10.5281/zenodo.17617792).

https://zenodo.org/records/17617792

## Data availability

All medical and marketing ratings necessary to replicate the results of this study are publicly available on Zenodo (doi: 10.5281/zenodo.17617792). The full dataset of generated model responses is available from the corresponding author upon reasonable request. No custom code was used in this study.

## Author Contributions

FR and CvE designed the study. FR analyzed data and wrote the original manuscript. LJB and CM rated datasets and reviewed the manuscript. CL and CvE supervised the study and reviewed the manuscript. All authors have approved the final manuscript.

## Acknowledgement

We wish to thank Katharina Beuchert, Paul Bienwald, Stephanie Klein, and Leonie Stroh for providing additional ratings for the case vignettes.

