## Supplementary Information for "Leveraging large language models to address common vaccination myths and misconceptions"

**Supplementary Table 1.** Overview of the Robert Koch Institute's “fact sandwiches” used as input for this study*.

| Topic | Myth | Fact |
| --- | --- | --- |
| Efficacy | “Vaccines are pointless as they do not provide 100% protection.” | “Vaccination is meaningful even though protection be 100 %.” |
| Risk-benefit analysis | “The vaccine is more dangerous than the infection.” | “The infection and its possible  complications are significantly more dangerous than the recommended vaccine.” |
| Childhood vaccination | “Infants and toddlers are being vaccinated too early.” | “Vaccinating infants and toddlers is  important for their health.” |
| Safety | “The side effects and risks of vaccines are incalculable.” | “The risks and side effects of vaccines are determined in clinical trials prior to their approval, and following approval they continue to be monitored worldwide.” |
| Safety | “Vaccinations cause the diseases that they are supposed to protect us against.” | “Vaccines cannot cause the disease they are meant to protect against.” |
| Safety | “The mRNA in vaccines changes the human genome – our DNA.” | “The mRNA in vaccines cannot be  incorporated into the genome of our cells.” |
| Safety | “Vaccines cause infertility.” | “Vaccines have no effect upon  fertility and are safe for those planning to have children.” |
| Safety | “Vaccines can cause cancer.” | “Vaccines can protect against cancer.” |
| Safety | “The measles, mumps, and rubella vaccine can cause autism.” | “The measles, mumps, and rubella vaccination can be ruled out as a possible cause of autism.” |
| Safety | “Vaccinations cause allergies.” | “Vaccines do not cause allergies.” |
| Communication | “The side effects of vaccinations are concealed.” | “All known side effects of vaccinations are communicated via different channels.” |

*Reference: https://www.rki.de/EN/Topics/Infectious-diseases/Immunisation/Information-material/Vaccination-myths/effectively-debunking-misinformation-node.html (accessed on July 1, 2025)

**Supplementary Table 2**. Model performance regarding communication clarity as rated by marketing experts. Smaller mean ranks indicate better performance (scale: 1 = best, 2 = intermediate, 3 = worst).

| Model (Vendor) | Mean Rank  (Standard Deviation) | | Median | Absolute Count Rank 1 | Relative Count Rank 1 |
| --- | --- | --- | --- | --- | --- |
| GPT-5 (OpenAI) | | 1.93 (0.084) | 2.0 | 30 | 34.1% |
| Gemini 2.5 Flash (Google) | | 1.83 (0.087) | 2.0 | 38 | 43.2% |
| Claude Sonnet 4 (Anthropic) | | 2.24 (0.086) | 2.0 | 20 | 22.7% |
